# Assessment of Prognostic Value of Cystic Features in Glioblastoma Relative to Sex and Treatment with Standard-of-Care

**DOI:** 10.1101/19013813

**Authors:** Lee Curtin, Paula Whitmire, Cassandra R. Rickertsen, Gina L. Mazza, Peter Canoll, Sandra K. Johnston, Maciej M. Mrugala, Kristin R. Swanson, Leland S. Hu

**Author notes:** **Corresponding Author:** Lee Curtin. first authors contributed equally (Lee Curtin, Paula Whitmire). co-senior authors (Kristin R. Swanson, Leland S. Hu). **Addresses:** LC, PW, CRR, MMM, KRS and LSH - 5777 E Mayo Blvd, Phoenix, AZ, 85054. PC - 630 West 168th Street, Mailbox 23, New York, NY 10032, GM – 13400 E Shea Blvd, Scottsdale, AZ, 85259. **Author Contributions:** LC and PW carried out the literature search, data analysis and manuscript preparation. CRR provided expertise in cystic GBM MRI presentation. GLM verified the statistical analyses and conclusions of this work. PC provided pathological expertise. SKJ was involved in data curation. MMM provided clinical and neuro-oncology expertise. KRS provided data, expertise, was involved in manuscript presentation and co-supervised this project. LSH provided the expertise of a neuro-radiologist, vetted data, helped in manuscript preparation and co-supervised this project. All authors have approved this manuscript for submission.

## Abstract

Glioblastoma (GBM) is the most aggressive primary brain tumor and can have cystic components, identifiable through magnetic resonance imaging (MRI). Previous studies suggest that cysts occur in 7-23% of GBMs and report mixed results regarding their prognostic impact. Using our retrospective cohort of 493 patients with first-diagnosis GBM, we carried out an exploratory analysis on this potential link between cystic GBM and survival. Using pretreatment MRIs, we manually identified 88 patients with GBM that had a significant cystic component at presentation and 405 patients that did not. Patients with cystic GBM had significantly longer overall survival and were significantly younger at presentation. Within patients who received the current standard of care (SOC) (N=184, 40 cystic), we did not observe a survival benefit of cystic GBM. Unexpectedly, we did not observe a significant survival benefit between this SOC cystic cohort and patients with cystic GBM diagnosed before the standard was established (N=40 with SOC, N=19 without SOC); this significant SOC benefit was clearly observed in patients with noncystic GBM (N=144 with SOC, N=111 without SOC). When stratified by sex, this significant survival benefit was only preserved in male patients (N=303, 47 cystic). We report differences in the absolute and relative sizes of imaging abnormalities on MRI and the prognostic implication of cysts based on sex. We discuss hypotheses for these differences, including the possibility that the presence of a cyst could indicate a less aggressive tumor.

## Introduction

Glioblastoma (GBM) is the most common and aggressive primary brain tumor with a median survival of only 15 months with current standard-of-care treatment(1). The hallmark of GBM on T1-weighted MRI with gadolinium contrast (T1Gd MRI) is a heterogeneously enhancing mass, often with a dominant central non-enhancing component, comprised of central necrosis. The margins of the central necrosis are characteristically irregular and ill-defined (**Figure 1A**). In a pretreatment setting, T2-weighted (T2) MRI shows edema and low tumor cell densities that typically surround the T1Gd MRI enhancing region.

**Fig. 1.**
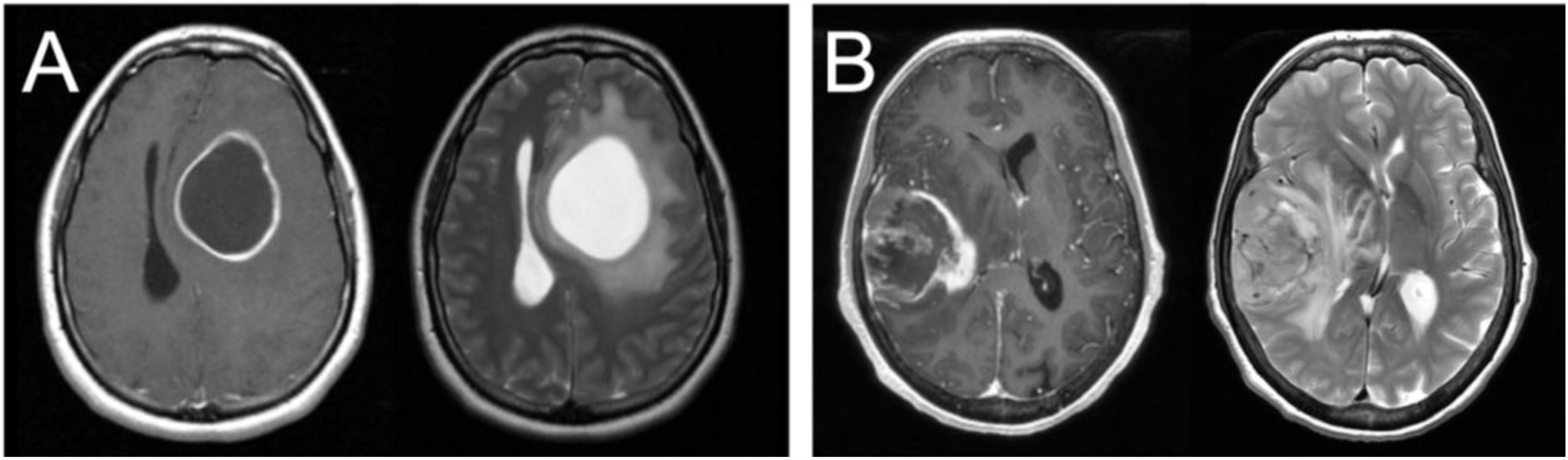
Example T1Gd and T2 MR images from cystic and noncystic GBM patients. **(A)** Cystic GBM. Cysts are identifiable by their round shape, thin contrast-enhancing rim on T1Gd (left), and their smooth homogenous appearance with clear borders on T2 (right) or T2-FLAIR **(B)** Noncystic GBM, note the nonuniform central hypointense region on T1Gd that does not show hyperintensity on T2.

By comparison, a percentage of GBM tumors present with one or more central cystic component(s) instead of or in tandem with necrosis. As opposed to irregular margins of central necrosis, cysts are often defined by smooth, thin, sharply defined margins of enhancement surrounding the non-enhancing cyst. Cystic fluid is typically isointense with cerebrospinal fluid (CSF) on T2 and FLAIR MRI and has a notably smooth texture. While these cysts are more common in lower grade gliomas (e.g., juvenile pilocytic astrocytoma, ganglioglioma) and non-glioma tumors with good prognosis (e.g., hemangioblastoma), between 7-23% of GBM tumors can also present with dominant cystic components (rather than necrosis) (**Figure 1B**)(2,3). We will refer to these as cystic GBMs and those without a significant cystic component as noncystic GBMs.

Current clinical practice does not consider the presence of a cyst when formulating treatment plans or prognosticating patients’ survival. There have been a number of studies that have assessed prognostic implications of cystic GBMs with inconsistent results. One retrospective review matched 22 patients with cystic GBM with 22 patients with noncystic GBM treated between 1993 and 2002. The authors found an insignificant trend for overall survival (p=0.1, log-rank test), and found that patients with cystic GBM had a significantly longer time to recurrence (2). Another study of 5 cystic GBMs and 32 noncystic GBMs treated between 1990 and 2004 also found that patients with cystic GBM had a longer time to recurrence and a median survival time of 19.8 months compared to only 12.8 months for noncystic GBM (4). This is supported by a smaller study of 7 cystic and 14 noncystic GBMs treated between 2000 and 2009 that found patients with cystic GBM lived significantly longer after surgery (5). Another recent study of patients with GBM treated between 2004 and 2014 found a survival benefit for those with cystic GBM (41 cystic, 139 noncystic) (3). A study published in 2016 looking into the benefits of maximizing extent of resection presented 118 GBM patients treated between 1993 and 2012 with a cyst on imaging (and 1108 without) and found this was beneficial to overall survival while accounting for age at surgery, preop KPS score, previous treatment, T1Gd enhancing volume and extent of resection (6). However, two of the larger studies to date found no survival benefit associated with cystic GBM presentation. One of these studies reviewed 354 GBM patients (37 cystic, 317 noncystic) treated consecutively between 2005 and 2009 at a single institution (7) and the other retrospectively reviewed 351 consecutive GBM patients (27 cystic, 324 noncystic) treated between 1997 and 2011 (8).

Given the higher prevalence of cysts in lower grade tumors, there have been a number of studies looking for a mechanism behind the improved prognosis of patients with cystic GBM, including notable molecular compounds present in cystic GBM fluid, histopathological differences between cystic and noncystic GBM and prevalence of IDH1 mutants in cystic GBM (9,10). Other studies saw histological differences between cystic and noncystic GBMs; cystic GBM had more well-defined tumor cell borders, suggesting that cystic GBM tumor cells are less migratory (2,4). Multiple studies have found that cystic GBM patients tend to be younger than noncystic patients (4,5,7). IDH1 mutation, considered a genetic marker for secondary GBM, was found only in 2/27 patients with cystic GBM in one study that concluded cystic GBM was unlikely to be progression from lower grade glioma (8).

The lack of consensus about the prognostic value and biological indications of cystic components at GBM presentation leaves uncertainty in the clinical interpretation of this phenomenon. Elucidating whether cysts provide a prognostic benefit and better understanding the biological mechanism behind this phenomenon will allow for more informed decision-making in these situations. In this investigation, we sought to determine whether cysts provide a prognostic benefit and identify other image-based and clinical characteristics associated with cystic GBM. Additionally, we looked for patient and tumor characteristics that could help explain the biological mechanism of the development of cysts in GBM. We further explore these effects within patient sex and with respect to standard-of-care treatment.

## Methods

### Patient Cohort

All patients from our in-house multi-institutional glioma repository that met the following criteria were included in this cohort: A) first-diagnosis GBM, B) available age at diagnosis, sex, and overall survival (confirmed death, alive, or lost to follow-up), and C) available pre-surgical T1Gd MR image. This resulted in a cohort of 493 newly-diagnosed GBM patients (190 females and 303 males). For the patients with this information available in our repository, extent of resection (N=350) and progression free survival were also collected (N=166). We also collected IDH1 and MGMT status when available (N=115 and N=89, respectively). Extent of resection was abstracted from clinical progress reports and radiological reports. Karnofsky performance statuses (KPS) were available in 135 patients. Since many of the patients in our cohort were diagnosed before the current standard of care (SOC) for newly-diagnosed GBM was established in 2005, the whole cohort did not receive the same SOC treatment. In order to assess the impact of cysts on patients receiving the current SOC, as established in 2005, we created a subset of patients who were known to receive maximal safe resection, concurrent XRT and TMZ, and at least one cycle of TMZ after diagnosis and called these patients “current SOC patients” (N=184). We isolated patients who were not known to receive current SOC and were known to have been diagnosed before the SOC was established in 2005 and called these patients “not current SOC patients” (N=130). For more information on the patient cohort, see **Table 1**. We present the top 10 most common known treatments (excluding surgery) received within our cohort in **Supplement 1**. Cohort matching was also carried out using propensity score matching using the matchit package in R(11) (further details in **Supplement 2**). All patients included in this investigation were consented prospectively or approved for retrospective research by Mayo Clinic Institutional Review Board.

**Table 1.** Patient and tumor characteristics of cystic and noncystic GBM. The means and medians in this table were reported to four significant figures.

|  | Cystic GBM (N=88) |  |  | Noncystic GBM (N=405) |  |  | Statistical Comparison |
| --- | --- | --- | --- | --- | --- | --- | --- |
|  | Median | Mean | N | Median | Mean | N |  |
| Age at Diagnosis (years) | 56 | 53.32 | 88 | 60 | 59.11 | 405 | **p=0.0016**<br>(t-test) |
| Sex |  |  | Female: 41<br>Male: 47 |  |  | Female: 149<br>Male: 256 | p=0.087<br>(chi-squared) |
| Necrosis/Cystic volume (mm<sup>3</sup>) | 14040 | 19510 | 87 | 4700 | 7898 | 405 | **p<0.0001**<br>(t-test) |
| T1Gd volume (mm<sup>3</sup>) | 46310 | 51020 | 88 | 25850 | 31140 | 405 | **p<0.0001**<br>(t-test) |
| T1Gd velocity (mm/day) | 0.02028 | 0.03024 | 14 | 0.08232 | 0.1149 | 54 | **p=0.039**<br>(t-test) |
| T2/FLAIR volume (mm<sup>3</sup>) | 104700 | 104500 | 59 | 85430 | 90900 | 267 | p=0.053<br>(t-test) |
| T1Gd/T2FLAIR | 0.5020 | 0.5038 | 59 | 0.3290 | 0.3860 | 267 | **p=0.00030**<br>(t-test) |
| Preop KPS | 90 | 86.67 | 33 | 80 | 82.33 | 102 | **p=0.021**<br>(t-test) |
| Extent of Resection |  |  | Biopsy: 7<br>STR: 33<br>GTR: 35 |  |  | Biopsy: 65<br>STR: 102<br>GTR: 108 | **p=0.025**<br>(chi-squared) |
| IDH1 Status |  |  | Wild Type: 32 (84%)<br>Mutant: 6 (16%) |  |  | Wild Type: 71 (92%)<br>Mutant: 6 (8%) | p=0.19<br>(chi-squared) |
| MGMT Status |  |  | Unmethylated: 10 (59%)<br>Methylated: 7 (41%) |  |  | Unmethylated: 50 (69%)<br>Methylated: 22 (31%) | p=0.40<br>(chi-squared) |
| Progression Free Survival (days) | 290 |  | 36 | 275 |  | 130 | p=0.97<br>(log-rank) |
| Overall Survival (days) | 680 |  | Deceased: 70<br>Alive/LTFU: 18 | 456 |  | Deceased: 348<br>Alive/LTFU: 57 | **p=0.0012**<br>(log-rank) |
| Overall Survival (Current SOC only) (days) | 750 |  | Deceased: 28<br>Alive/LTFU: 12 | 678 |  | Deceased: 113<br>Alive/LTFU: 31 | p=0.29<br>(log-rank) |

### MR Evaluation

The tumor-associated imaging abnormality on patients’ pre-surgical MR images were segmented by trained individuals. From these segmentations, we calculate the volume of the imaging abnormality (mm3). Every patient in this cohort has a “T1Gd volume”, quantifying the size of the imaging abnormality on the pre-surgical T1Gd image that was closest to the date of surgery. “T1Gd volume” includes both the central hypointense region of necrosis or cystic fluid and the surrounding contrast-enhancing region. The central hypointense area of necrosis or cystic fluid is also segmented separately and used to calculate a “necrosis/cystic volume”. Due to the varied imaging protocols between patients, the accuracy of volumes presented in this work will also vary between patients. For patients that had multiple pre-treatment T1Gd images (in a 10+ day interval), we took the T1Gd volume of both images, converted them to radii using a spherically-equivalent volume, and calculated a “T1Gd radial growth velocity” (mm/day) (N=68). Most patients also had an available pre-surgical T2-FLAIR MRI (or T2 MR image if T2-FLAIR was not available) (N=326), which we used to calculated a “T2/FLAIR volume”.

### Identification of Cysts

All cystic GBM and noncystic GBM patients were identified using pre-surgical MR images by two trained individuals (L.C. and P.W.) and a randomly chosen subset (consisting of 20 cystic and 20 noncystic GBM MRIs) was vetted by a neuroradiologist (L.S.H). Cysts were defined by smooth, thin, sharply defined margins of enhancement surrounding the periphery of the non-enhancing cyst. Where available, the T2/FLAIR MRI region corresponding to the hypointensity on T1Gd was clearly defined, consistent with CSF, and smooth in texture. In order to be deemed a “cystic GBM”, the cyst(s) needed to be comparable in size or larger than the rest of the tumor-related non-enhancing abnormality. GBM tumors that did not fit this definition were deemed as “noncystic GBM”. Examples of T1Gd and T2 MRIs for both a cystic and noncystic GBM are shown in **Figure 1**.

### Statistical analyses

Kaplan-Meier curves were used to visualize differences in survival between two groups and log-rank tests were used to assess the significance of these differences. Cox proportional hazards models (CPH), both univariate and multivariate, were also used to assess the impact of patient variables on overall survival. Two-sided independent t-tests with Welch’s corrections were used to test whether the means of a particular variable were statistically different between two groups. All statistical tests and figures were generated using R(12) and the packages survminer(13), ggplot2(14), and survival(15). In order to assess the role of sex differences in this investigation, all tests were also performed separately on male and female patients. Any of the results from this sex differences analysis that are not reported in the main text are listed in the supplement. Additionally, the age-adjusted rate of the formation of cysts in newly-diagnosed GBM was calculated using the CBTRUS 2011-2015 GBM population(16) as the “standard population”. For each age group, age-adjusted rate of cyst is calculated as (cysts in our cohort in that age group)/(number of GBM patients in CBTRUS data in that age group). The resulting “rate” is not intended for prediction of cyst prevalence, but to allow for the comparison of cyst frequency across age groups in a way that normalizes for the frequency of GBM in that age group (**Supplement 3**).

### Ethical Approval

All procedures performed in the studies involving human participants were in accordance with the ethical standards of the institutional and/or national research committee and with the 1964 Helsinki declaration and its later amendments or comparable ethical standards. Our de-identified data repository of patients with brain cancer includes retrospective data collected from medical records and prospective data collection. Research conduct on the data repository is approved by Mayo Clinic Institutional Review Board (17-009688). Prior to collection of retrospective data informed consent was waived for those participants by the Mayo Clinic Institutional Review Board (IRB# 15-002337). Written informed consent was obtained for all prospectively enrolled participants as approved by Mayo Clinic Institutional Review Board (IRB# 17-009682).

### Data Sharing

The data used in this study are part of a larger collection curated by Kristin R. Swanson. These data contain protected health information and are therefore subject to HIPAA regulations. If requested, data may be made available for sharing to qualified parties as soon as is reasonably possible, so long as such a request does not compromise intellectual property interests, interfere with publication, invade subject privacy, or betray confidentiality. Typically, data access will occur through a collaboration and may require interested parties to obtain an affiliate appointment with the author’s institution. Data that are shared will include standards and notations needed to interpret the data, following commonly accepted practices in the field.

## Results

### Patients with cystic GBM have significantly longer overall survival among patients receiving all treatments, but not among current SOC patients

In our patient cohort, cystic GBM patients (N=88) have a significantly longer overall survival (OS) than noncystic patients (N=405) (log-rank, p=0.0012), but there is no significant survival difference among patients that received the current standard of care (p=0.29, N=184) (**Figure 2**). When controlling for age as a prognostic factor among all patients (N=493), the existence of cyst remained significant in predicting survival in a multivariate CPH analysis (HR = 0.753, p=0.032). When controlling for preoperative KPS score (N=135), the existence of cyst did not remain significant in multivariate CPH analysis (HR = 0.8042, p=0.321). Among all patients with known extent of resection (N=350), we performed a multivariate CPH controlling for cyst status, age, extent of resection and treatment with current SOC. In this setting, we found that the presence of cyst approached significance (Cystic: yes = 1) (HR=0.751, p=0.052), independent of other variables like age at diagnosis (HR=1.03, p<0.0001), extent of resection (biopsy = 1, STR = 2, GTR = 3) (HR=0.681, p<0.0001), and treatment protocol (Current SOC: yes = 1) (HR=0.553, p<0.0001). Comparing 5-year survival rates between cystic and noncystic GBM yields significant differences (chi-squared p=0.011). We see a 20% 5-year survival rate for cystic GBM patients (N=18 of 88) and an 11% equivalent in the noncystic GBM cohort (N=43 of 405). We do not see the same significance within the SOC patients (chi-squared p=0.67). The 5-year survival rate for cystic GBM receiving SOC is 23% (N=9 of 40), while the corresponding noncystic GBM rate is 19% (N=28 of 144).

**Fig. 2.**
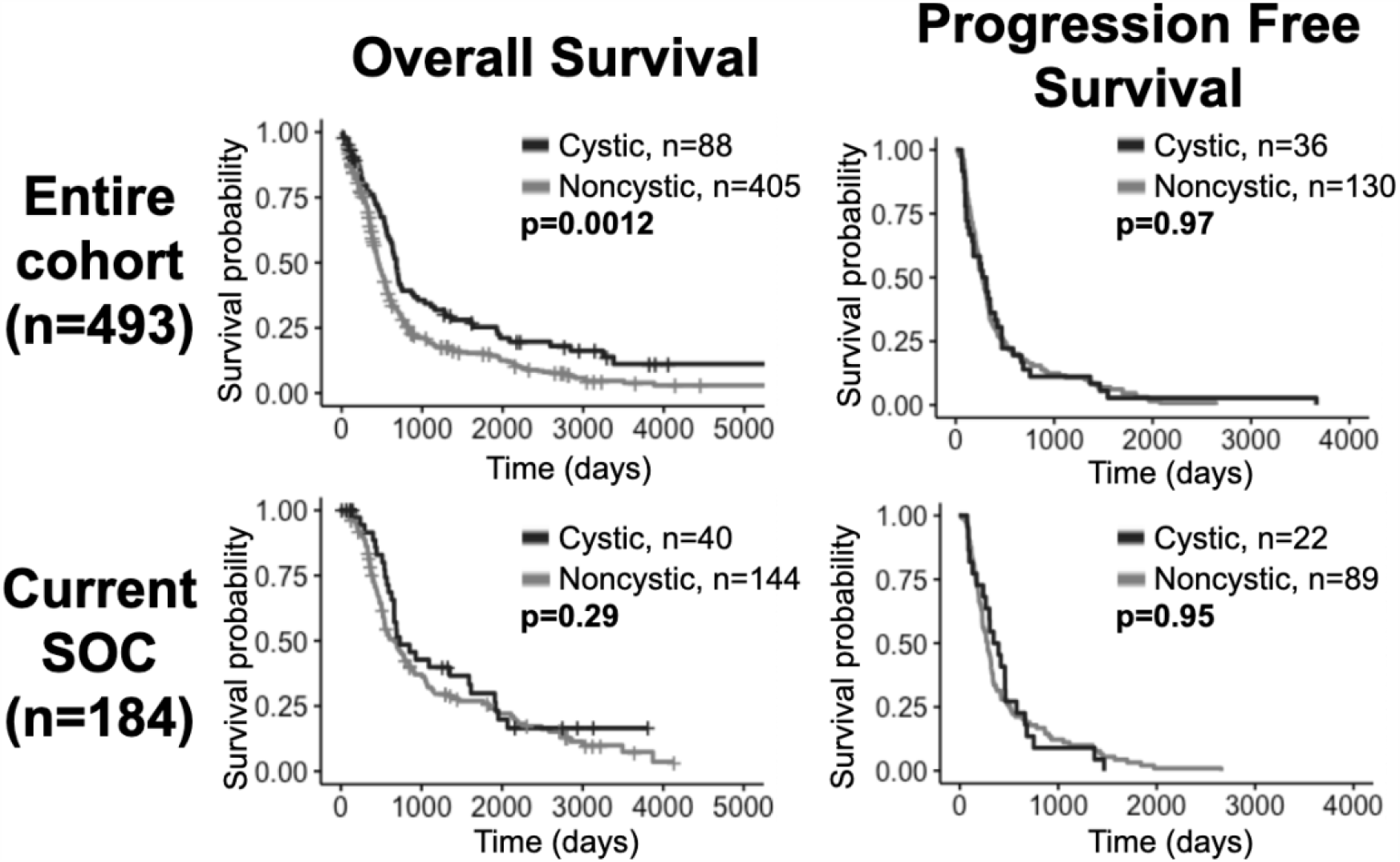
Overall survival benefit for cystic GBM but no significant differences in progression free survival. Cystic GBM have longer survival among the entire cohort, but not among current SOC patients. Comparisons of OS (left column) and progression free survival (right column) of cystic versus noncystic GBM patients. The top row shows the comparisons among all patients and the bottom row shows the comparisons among current SOC patients only. While cystic patients have an OS benefit compared to noncystic patients when looking at the whole cohort, this difference is not statistically significant among current SOC patients.

Matching noncystic GBM patients with cystic GBM patients by the top 10 most common known treatments (found in **Supplement 1**) preserved significant OS differences in 88.8% of tested cases (1000 unique matches, mean p=0.024, N=88), see **Supplement 2**. This result was weakened to 48.2% of tested cases with the inclusion of matched EOR in a reduced cohort (1000 unique matches, mean p=0.067, N=75). Significant OS differences are still observed while matching a subset of patients (N=59) by SOC status, age at diagnosis, sex and the top 10 most common known treatments received (2 unique matched cohorts, both p=0.012), see **Supplement 2**. This result was no longer significant with the inclusion of EOR (1 unique matched cohort, p=0.35, N=50).

### Patients with cystic GBM are younger, tend to have slower tumor growth velocity, and have relatively less T2/FLAIR abnormality

We present a summary of our comparisons between cystic GBM and noncystic GBM patients in **Table 1**. Cystic GBM patients are significantly younger than noncystic GBM patients at the time of diagnosis (t-test, p=0.0016) (**Figure 3A**). We calculated an age-adjusted rate of cystic presentation in GBM using the CBTRUS 2011-2015 population as our “standard population” of GBMs, which allows us to normalize the frequency of cystic presentation in a particular age group to the frequency of GBM diagnosis in that age group (more details on this method in **Supplement 3**). This calculation showed that cystic GBM is much more common among younger patients, particularly among patients 20-34 years old, and were least common among patients 75-84 years old (**Figure 4**). We also note that patients with cystic GBM had a significantly higher KPS than patients with noncystic GBM (t-test p=0.02), see **Table 1**.

**Fig. 3.**
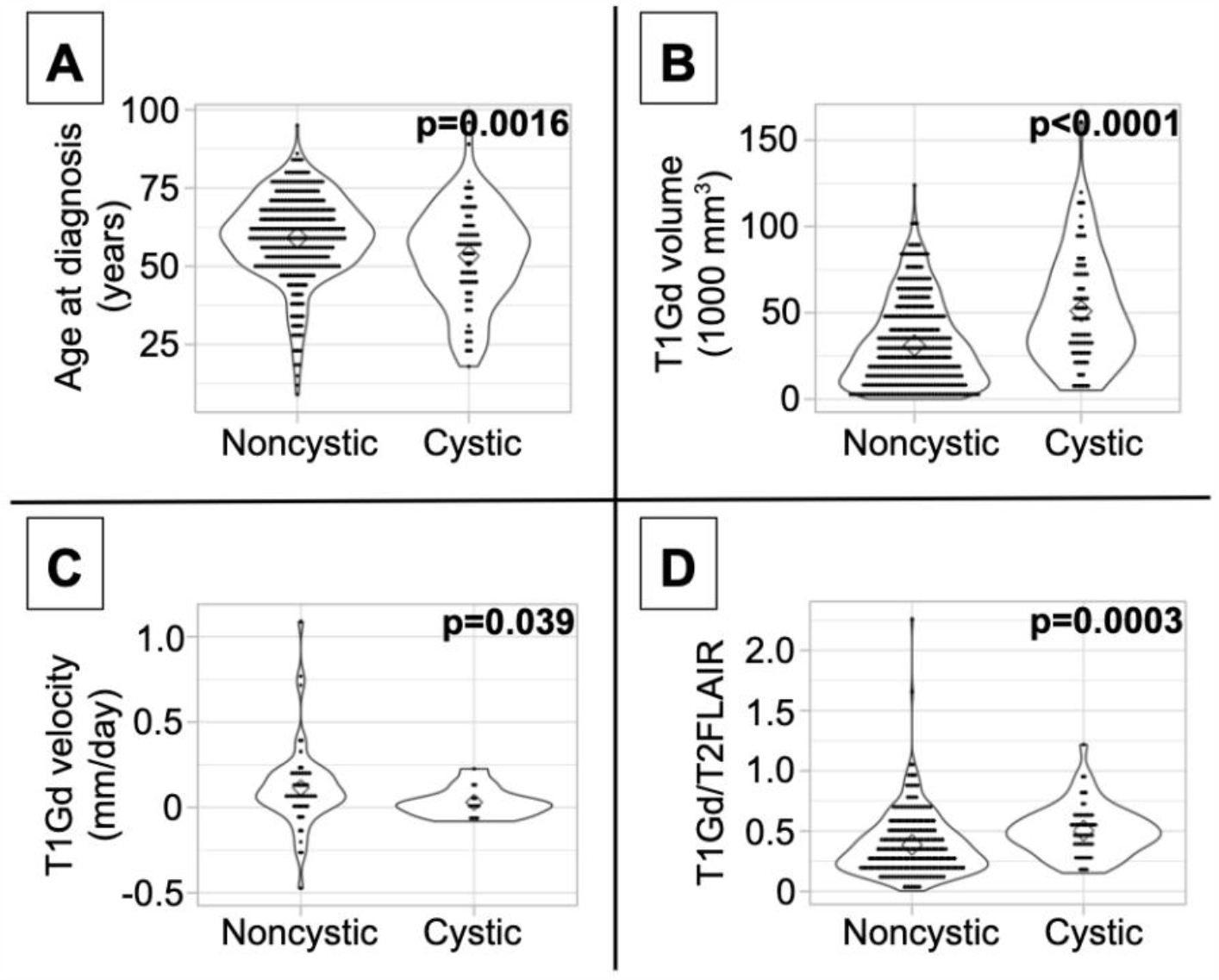
Differences in imaging abnormalities and age at diagnosis. Compared to noncystic GBM patients, **(A)** cystic patients were younger, **(B)** had larger tumor volume on T1Gd MRI, **(C)** had slower pre-surgical growth velocity on T1Gd MRI, and **(D)** had less volume on T2/FLAIR MRI relative to volume on T1Gd. We present the results of these tests amongst patients known to have received the current SOC in **Supplement 5**.

**Fig. 4.**
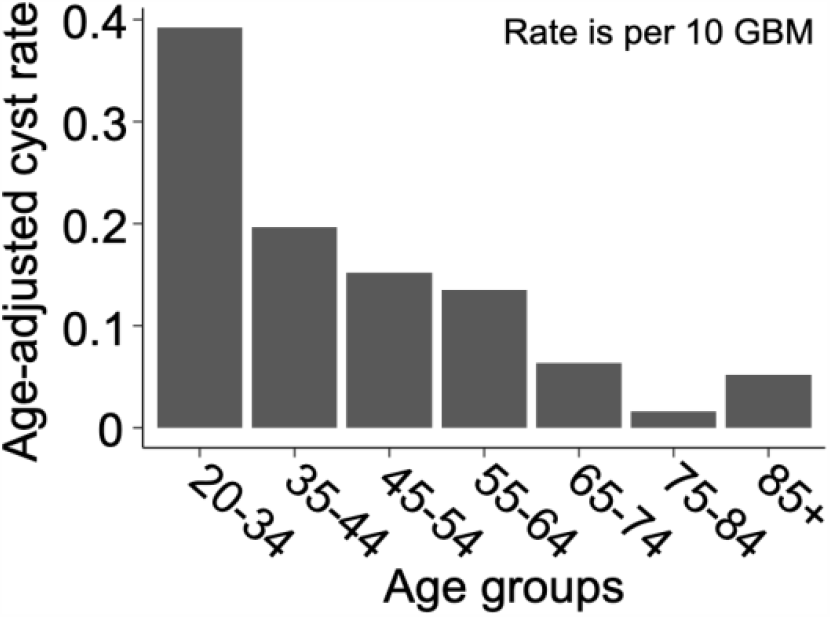
Cystic GBM is more common in younger patients. Cysts are more common amongst younger GBM patients. Age-adjusted rate of presentation with cyst among GBM patients using CBTRUS 2011-2015 GBM population as the “standard population.” Age-adjusted rates are given as cystic cases per 10 GBM cases. The resulting “rate” is not intended for prediction of cyst prevalence, but to allow for the comparison of cyst frequency across age groups in a way that normalizes for the frequency of GBM in that age group. Comparing the rates across age groups shows that cystic GBM is most common among 21-34 year-olds and least common among 75-84 year-olds.

Using tumor volume data from multi-modal MR images, we found that cysts have larger volumes of T1Gd abnormality (p<0.0001) (**Figure 3B**) and insignificantly tend to have larger volumes of T2/FLAIR abnormality (p=0.053) (**Supplement 4**). This was expected since cysts tend to be larger than necrosis at presentation, supported by the larger necrosis/cyst volume in cystic patients (p<0.0001) (**Supplement 4**), and the cystic volume is a component of the T1Gd and T2/FLAIR abnormalities. When comparing the relative sizes of the T1Gd and T2/FLAIR volumes (T1Gd/T2FLAIR), we found that cystic GBM have a smaller relative T2/FLAIR component compared to noncystic GBM (p=0.0003) (**Figure 3D**). Cystic GBM patients also have a significantly slower (p=0.039) pre-surgical T1Gd growth velocity compared to noncystic GBM patients (**Figure 3C**).

### Patients with cystic GBM do not significantly benefit from current SOC, while those with noncystic GBM show the expected survival benefit

When comparing cystic current SOC patients to cystic not current SOC patients, we surprisingly do not see significant OS differences (log-rank, p=0.28 **Figure 5A**). As expected we see a very significant OS benefit in the analogous result for noncystic GBM patients (p<0.0001, **Figure 5B**). These two results hold amongst patients with known IDH1 wild type status. Patients with cystic IDH1 wild type did not show a survival benefit from the current SOC (p=0.99, **Figure 5C**) whereas for noncystic IDH1 wild type there is a clear benefit of current SOC (p<0.0001 **Figure 5D**). Among current SOC patients, cystic patients did not have significantly improved OS compared to noncystic patients (p=0.29) (**Figure 2**), but among not current SOC patients, cystic patients did have significantly longer OS compared to noncystic patients (p=0.049) (**Supplement 6**). We observed the same significance trends in patients with known IDH1 wild type GBM. Cystic GBM IDH1 wild type patients who did not receive the current SOC received a survival benefit compared to noncystic GBM IDH1 wild type patients (p=0.029) (**Supplement 6**).

**Fig. 5.**
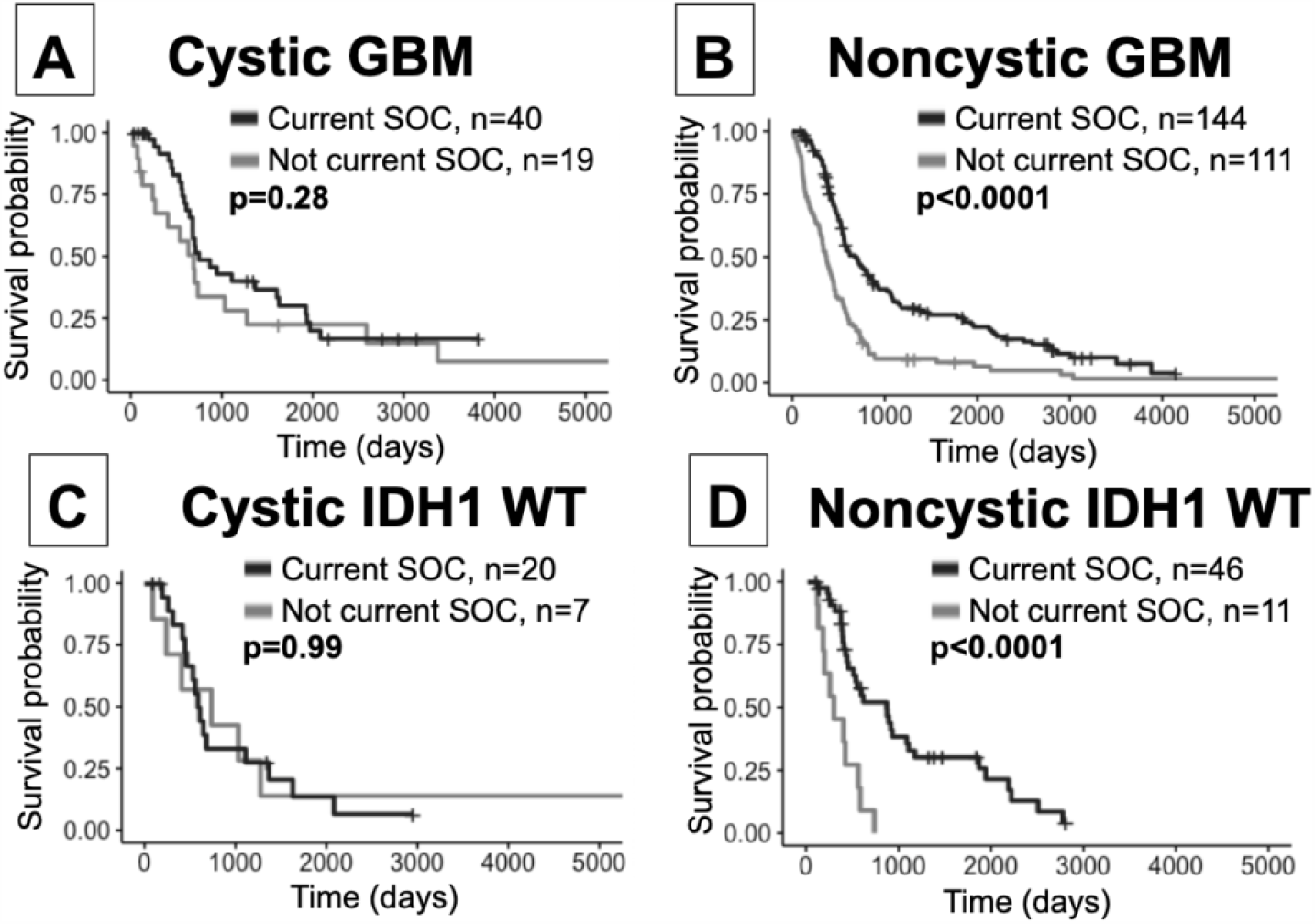
Current SOC benefit is enriched in noncystic GBM compared with cystic GBM. **(A)** Cystic GBM current SOC patients (median OS: 750 days) did not have significantly improved survival compared to cystic GBM not current SOC patients (median OS: 691 days). **(B)** Noncystic current SOC patients (median OS: 678 days) do show an expected survival benefit compared to noncystic not current SOC patients (median OS: 370 days). **(C)** Cystic current SOC patients (median OS: 737 days) with known IDH1 wild type do not show a significantly improved survival compared to not current SOC IDH1 wild type cystic GBM patients (median OS: 600 days). **(D)** Noncystic IDH1 wild type current SOC patients (median OS: 858 days) have a large survival benefit compared to IDH1 wild type noncystic not current SOC patients (median OS: 286 days).

### Female patients with cystic GBM have less of a prognostic benefit compared to males

When males and females were combined, cystic patients had a survival benefit compared to noncystic patients among patients receiving all treatments, but this difference was not significant among current SOC patients. In order to identify potential sex differences, all of the tests in this investigation were also run on male and female cohorts separately (**Supplement 7**). Among males, the survival difference between cystic and noncystic patients is significant among patients with all treatments (log-rank, p=0.0006), and is insignificant but visually trending towards significance among current SOC patients (p=0.095). Among females, there was no significant survival difference between cystic and noncystic patients among patients receiving all treatments (p=0.29), but cystic patients do have improved median OS (cystic median OS: 589 days, noncystic: 454 days). Among female current SOC patients, we did not observe a survival difference (p=0.63) and unexpectedly the noncystic patients had longer median OS than the cystic patients (cystic median OS: 681 days, noncystic: 858 days) (**Figure 6**). Additionally, when males and females were combined, cystic patients had relatively less T2/FLAIR volume compared to noncystic (higher T1Gd/T2FLAIR ratio, cystic mean: 0.504, noncystic mean: 0.386, p=0.0003). When the patients are separated by sex, this difference in relative volume was more notable among females (T1Gd/FLAIR ratio, cystic mean: 0.555, noncystic mean: 0.397, p=0.006) than it was among males (T1Gd/FLAIR ratio, cystic mean: 0.461, noncystic mean: 0.380, p=0.027) (**Supplement 7**). The female cystic ratio was not significantly different than the male cystic ratio (p=0.091). In multivariate CPH analyses within patient sex, the presence of cystic GBM was significantly beneficial to males (p=0.027, HR=0.655) while accounting for age at diagnosis but was not significantly beneficial to females (p=0.461, HR=0.866) (**Supplement 7**).

**Fig. 6.**
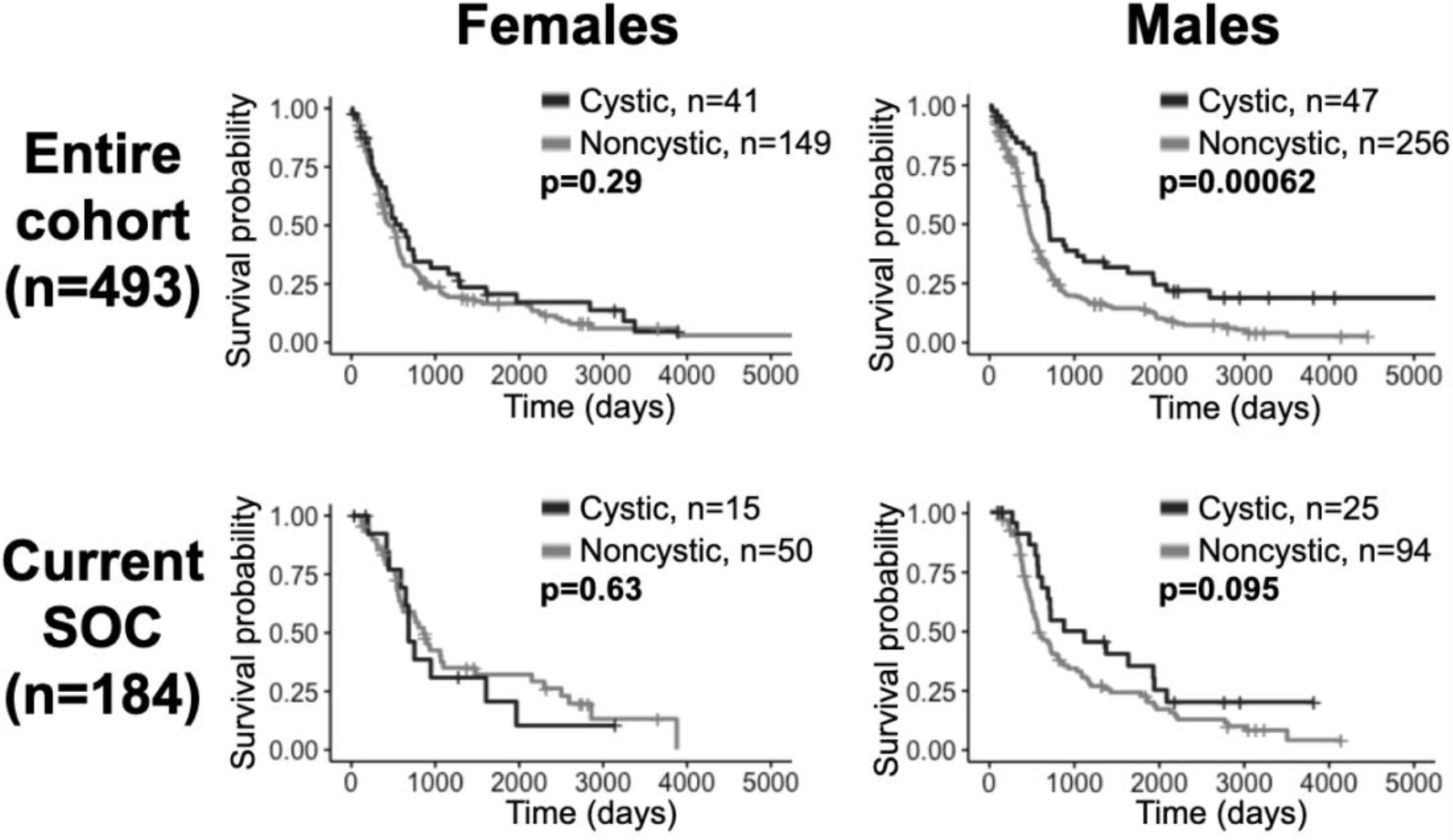
Cystic GBM survival benefit is driven by males. Overall survival comparisons between cystic and noncystic patients, both among all patients (top row) and current SOC patients (bottom row), separated into females (left column) and males (right column). Cystic GBM males had improved survival compared to noncystic GBM males among the entire cohort and they tended to have better outcomes among the current SOC patients. Among females, cystic GBM did not have significantly longer overall survival compared to noncystic GBM.

Among the patients with available histological information, we did not observe a difference in the prevalence of IDH1 mutation or MGMT methylation between cystic and noncystic patients (**Table 1**). 115 patients had IDH1 mutation status available, consisting of 103 IDH1 wild-type patients (32 cystic and 71 noncystic) and 12 IDH1 mutant patients (6 cystic and 6 noncystic) (Fisher’s exact test, p=0.21). 89 patients had available MGMT methylation status, including 29 unmethylated patients (7 cystic and 22 noncystic) and 60 unmethylated patients (10 cystic and 50 noncystic) (Fisher’s exact test, p=0.40). We observed survival results of patients with known IDH1 WT were consistent with observations regarding current SOC in our whole cohort (see **Figure 5** and **Supplement 5**). Patients with particular GBM MGMT status did not yield significant survival differences, which could be a result of low sample size.

Additionally, 50 of the 88 cystic GBM patients from our cohort had detailed notes from the diagnosing pathologist available for review. While all cystic patients in our cohort were diagnosed with grade IV glioblastoma by a pathologist, we reviewed these notes for evidence of unique pathological features in these tumors. We found that four cystic patients had features of giant cell GBM, three were diagnosed as gliosarcoma, and three showed evidence of developing from lower grade glioma. Two patients were observed to have oligodendroglial components and one patient had several characteristics that were suggestive of astroblastoma.

## Discussion

Collectively, previous studies have shown mixed results on the OS benefit of cystic GBM against noncystic GBM patients. The majority of studies observing an OS benefit for cystic GBM contain patients diagnosed before the current SOC (2,4–6,17). As described above, we have shown a significant increase in OS for cystic GBM patients when compared to noncystic GBM patients in a cohort of 493 patients (88 cystic GBM). However, the same analyses on only current SOC patients does not show the same significant result for OS. Similarly, the 5 -year survival rate is significantly higher for patients with cystic GBM within the whole cohort but not amongst current SOC patients. These results could be interpreted to suggest that the current SOC protocol affects noncystic GBM patients differently than cystic GBM patients. Indeed, we note an expected significant increase in OS from the current SOC within noncystic GBM patients, however, we find a non-significant increase in OS for cystic GBM patients when comparing current SOC patients to not current SOC patients (**Figure 5**). These differences are also preserved amongst patients with known IDH1 wild type GBM. This work naturally brings into question whether the potential underlying biological differences between cystic and noncystic GBM lead to a difference in responsiveness to current SOC therapies.

A separate study has analyzed the fluid in cystic GBM for various nutrients(9). The authors noted a variety of nutrients were present in cystic GBM fluid at similar or higher levels than in CSF. Particularly, glutamate, lactate and inorganic phosphate, which were significantly higher than in CSF, were all found in cystic GBM fluid and have been shown previously to act in a similar manner to growth factors for tumor cells (18–23). These results suggest that cystic fluid could act as a nutrient resource for GBM. If this is the case, then a nutrient-rich center of a GBM that is otherwise palisading necrosis could give a very different microenvironment to the tumor cells. Other papers have shown histological differences between cystic and noncystic GBM tumor cells; cystic tumor cells were less invasive into the brain parenchyma. This lack of invasion could lead to a better prognosis, tumor cells are less likely to migrate into other regions of the brain and lead to a less aggressive tumor overall. Another study found that brain tumor cystic fluid had an immunosuppressive effect on cultured lymphocytes (10).

Interestingly, presentation with a cyst did not seem to have the same prognostic value among female patients as it did among male (**Figure 6**). We also observed that cystic GBM had a more equal distribution of males and females (1.15:1 M:F ratio), while the distribution in noncystic GBM had considerably more males (1.72:1 M:F). Previous studies on sex differences in GBM have found differences in the impact of tumor invasion on outcome (24), but our literature search did not find any studies that looked at sex differences in cystic GBM.

We have shown using T1Gd growth velocity that cystic GBM tumors tend to grow slower than noncystic GBM tumors in our patient cohort. The observed slower growth for cystic GBMs could be indicative of a less aggressive tumor. Other studies have suggested that a GBM presenting with a cystic component could be an indication that the GBM is secondary, in that it developed from a low-grade glioma. There have been multiple papers with differing results on the prevalence of IDH1 mutations in cystic GBM(3,8). We did not find a significant IDH1 mutation prevalence in our cystic GBM cohort (**Table 1**) and observed survival differences amongst known IDH1 wild type cystic and noncystic GBM (**Figure 5** and **Supplement 5**). This suggests that our survival results are not a byproduct of IDH1 status.

From review of the available pathology reports for cystic GBM (50 out of 88), we found evidence that a few exhibited features of giant cell GBM, gliosarcoma, GBM with oligodendroglial components, and GBM that were suspected to evolve from lower grade gliomas; these features might be indicative of slower growing and/or le ss aggressive tumors and may be a contributing factor in our observed survival results. Another possible explanation for our observed OS benefit is that cysts lead to earlier symptom presentation in the course of the disease; this could be a result of pressure in the brain caused by the cyst, which would otherwise be solely caused by a tumor mass.

The precise biological mechanism of cyst formation in GBM is unclear. However, there is a consensus in the literature that edema acts as a precursor to cysts in both glioma and other tumors of the central nervous system (25,26). Vascular endothelial growth factor (VEGF) is known to be upregulated in GBM and plays a role in the breakdown of the blood brain barrier (27,28). VEGF has been shown to be expressed at high rates in GBM cyst fluid, which could be a contributing factor in GBM cyst formation (29,30). VEGFR pathway upregulation has been observed in rim-enhancing GBM (31). VEGF has also been linked to the presence of edema and cysts in other CNS tumors (32). A two-hit mechanism has been proposed for cyst formation in glioma: edema occurs as fluid extravasates from the leaky blood brain barrier at a faster rate than it is reabsorbed, and microcysts form due to microvascular phenomena which lead to a macroscale cyst over time (25).

Another noteworthy observation of this paper is the significantly younger age at presentation of patients with cystic GBM. This is not simply due to the earlier presentation of cystic GBM, as we have shown through an age adjusted rate in **Figure 4**. While the median age at diagnosis of GBM patients is about 65 years old and the age-adjusted incidence rate of GBM is highest among people 75-84 years old(16), we used age-adjusted rates to observe that cystic presentation was most frequent among GBM patients ages 20-34. Other papers have also found that cystic GBM patients tend to be younger than their noncystic counterparts (4,5,7). We also note that cystic GBM remained as a significant prognostic indicator when accounting for age in a multivariate Cox proportional hazard model.

### Limitations and future work

Although the patient cohort presented in this work is large, the findings of this study are caveated by some inherent limitations. This study is retrospective with incomplete records. Retrospective cohorts can generally bring the potential for retrospective biases, such as patient selection bias. Additionally, while we collect as much information as possible on our patients, we do not have complete molecular testing or clinical records for all patients. This patient cohort was diagnosed between 1993 and 2016, which covers a time period of a variety of treatment schedules as well as the introduction of a new treatment paradigm; this is inherent to any study of this nature but can introduce unexpected variability into our results. Keeping this in mind, we have collected available treatment information for the patients presented in this work and do not see significant differences of known received treatments (other than EOR) between patients with cystic GBM and those with noncystic GBM (see **Supplement 1**). We also presented overall survival results of matched cohorts (see **Supplement** 2). It is important also to note that results leading to a lack of statistical significance could be a result of a lack of power due to low sample size. A prospective study with more patients would be better suited to provide conclusive evidence on the survival benefit of cystic GBM patients, although it would be a significant challenge to find a prospective cohort of patients who are not receiving the current SOC. Such a study could take image-localized biopsies from cystic and noncystic GBM patients and determine corresponding histopathological, RNA sequencing, and genome sequencing analyses on the collected tissue samples. These would provide further evidence of potential biological differences in cystic and noncystic GBM tumors.

### Conclusion

While there has been previous evidence that cysts are a relevant prognostic indicator, some more recent publications have not found this to be the case. We find that cystic GBM is significantly beneficial to survival but do not see the same trend in patients receiving the current SOC; this could be due to a larger survival benefit of the current SOC to patients with noncystic GBM. Cysts seem to be indicative of less aggressive tumors that may preferentially benefit male patients.

## Data Availability

The data used in this study are part of a larger collection curated by one of the authors. These data contain protected health information and are therefore subject to HIPAA regulations. If requested, data may be made available for sharing to qualified parties as soon as is reasonably possible, so long as such a request does not compromise intellectual property interests, interfere with publication, invade subject privacy, or betray confidentiality. Typically, data access will occur through a collaboration and may require interested parties to obtain an affiliate appointment with the author's institution. Data that are shared will include standards and notations needed to interpret the data, following commonly accepted practices in the field.

## Acknowledgements

The authors gratefully acknowledge funding that made this research possible from the National Institutes of Health (R01NS060752, R01CA164371, U54CA143970, U54CA193489, U01CA220378, U54CA210180), the Arizona Biomedical Research Commission (ADHS16-162514), the James S. McDonnell Foundation (220020400TT), and the Ben and Catherine Ivy Foundation. This manuscript has been released as a pre-print at medRxiv (33). The authors thank Gustavo De Leon for useful discussions regarding the presentation of cysts on MR imaging, Harriet Crossland for discussions regarding the formation of cysts, and the Image Analysis Team at the Precision Neurotherapeutics Innovation Program for their contribution. Lastly, we honor the memory of the person who inspired this project, Anne Baldock - her passion and drive were taken from us too soon by a drunk driver. You are sorely missed, Anne.

